# Prediction of Complicated Ovarian Hyperstimulation Syndrome in Assisted Reproductive Treatment Through Artificial Intelligence

**DOI:** 10.1101/2024.04.17.24305980

**Authors:** Arash Ziaee, Hamed Khosravi, Tahereh Sadeghi, Imtiaz Ahmed, Maliheh Mahmoudinia

**Affiliations:** Medical Student, Student Research Committee, Faculty of Medicine, Mashhad University of Medical Sciences, Mashhad, Iran; Department of Industrial & Management Systems Engineering, West Virginia University, Morgantown, WV, US; Assistant Professor of Nursing, Department of Pediatrics, School of Nursing and Midwifery, Nursing and Midwifery Care Research Center, Akbar Hospital, Mashhad University of Medical Sciences, Mashhad, Iran; Assistant Professor, Department of Industrial & Management Systems Engineering, West Virginia University, Morgantown, WV, US; Assistant Professor of Obstetrics & Gynecology, Fellowship of Infertility, Supporting the Family and the Youth of Population Research Core, Department of Obstetrics and Gynecology, Faculty of Medicine, Mashhad University of Medical Sciences, Mashhad, Iran

**Keywords:** Ovarian Hyperstimulation Syndrome, Machine Learning, Assisted Reproductive Technology, In Vitro Fertilization, Data Augmentation

## Abstract

**Background:** This study explores the utility of machine learning (ML) models in predicting complicated Ovarian Hyperstimulation Syndrome (OHSS) in patients undergoing infertility treatments, addressing the challenge posed by highly imbalanced datasets.

**Objective:** This research fills the existing void by introducing a detailed structure for crafting diverse machine learning models and enhancing data augmentation methods to predict complicated OHSS effectively. Importantly, the research also concentrates on pinpointing critical elements that affect OHSS.

**Method:** This retrospective study employed a ML framework to predict complicated OHSS in patients undergoing infertility treatment. The dataset included various patient characteristics, treatment details, ovarian response variables, oocyte quality indicators, embryonic development metrics, sperm quality assessments, and treatment specifics. The target variable was OHSS, categorized as painless, mild, moderate, or severe. The ML framework incorporated Ray Tune for hyperparameter tuning and SMOTE-variants for addressing data imbalance. Multiple ML models were applied, including Decision Trees, Logistic Regression, SVM, XGBoost, LightGBM, Ridge Regression, KNN, and SGD. The models were integrated into a voting classifier, and the optimization process was conducted. The SHAP package was used to interpret model outcomes and feature contributions.

**Results:** The best model incorporated IPADE-ID augmentation along with an ensemble of classifiers (SGDClassifier, SVC, RidgeClassifier), reaching a recall of 0.9 for predicting OHSS occurrence and an accuracy of 0.76. SHAP analysis identified key factors: GnRH antagonist use, longer stimulation, female infertility factors, irregular menses, higher weight, hCG triggers, and, notably, higher number of embryos.

**Conclusion:** This novel study demonstrates ML’s potential for predicting complicated OHSS. The optimized model provides insights into contributory factors, challenging certain conventional assumptions. The findings highlight the importance of considering patient-specific factors and treatment details in OHSS risk assessment.

## INTRODUCTION

Ovarian hyperstimulation syndrome (OHSS) represents a critical challenge in assisted reproductive technology (ART), marked by a broad spectrum of clinical manifestations. This iatrogenic condition, typically occurring during the early luteal phase or early pregnancy following ovulation induction (OI) or ovarian stimulation (OS), varies in prevalence across different regions. In the United States, the incidence of moderate to severe OHSS in ART treatments was reported at about 1.2% in 2006, decreasing to 0.5% in 2014[1]. As per the European In Vitro Fertilization (IVF) -monitoring (EIM) Consortium, European figures show an incidence rate of only 0.3%[2].

OHSS is characterized by bilateral cystic enlargement of highly luteinized ovaries, leading to complications like vascular hyperpermeability and hemorrhagic ovarian cysts. Clinically significant OHSS occurs in 2-3% of cases, with milder forms affecting up to 20-30% of IVF patients[3]. Moderate OHSS can lead to symptoms like abdominal distention and nausea, with about 1.9% of patients needing hospitalization for severe effects, including hepatorenal failure and thromboembolism[4, 5]

OHSS typically occurs after ovarian stimulation with gonadotropins, causing an excessive ovarian reaction, including multiple follicle growth, high estradiol levels, and ovarian swelling. Exogenous hCG is crucial for oocyte maturation’s final steps. Its pathophysiology involves granulosa/luteal cells releasing vasoactive substances, leading to increased vascular permeability. Pregnancy following ovarian stimulation worsens OHSS as placental hCG boosts ovarian VEGF secretion, aggravating symptoms.[6–8].

Amid these challenges, artificial intelligence (AI) and machine learning (ML) have emerged as promising tools for improving OHSS management. Clinicians have traditionally relied on their expertise to prescribe the appropriate FSH dosage for follicular stimulation. The field of ML in OHSS management has seen significant advancements, such as optimizing the initial FSH dose in controlled ovarian hyperstimulation[9] and enhancing IVF protocols by predicting the effectiveness of each day of controlled ovarian stimulation (COS) monitoring[10]. These developments focus on predictive accuracy and prioritize patient safety and treatment efficacy. These models use historical data and predictive analytics to improve treatment outcomes, personalize therapies, and minimize the incidence of OHSS, representing notable progress in personalized medicine.

However, the challenge of imbalanced data in this field presents a significant obstacle in developing ML models specifically tailored to predict complicated OHSS occurrence and identify its essential contributing factors. This study addresses this gap by proposing a comprehensive framework for developing various ML models and data augmentation techniques to tackle this challenge. Notably, this study also focuses on identifying key factors influencing OHSS, thereby contributing valuable insights to the field and aiding in more effective complicated OHSS prediction and management.

## METHODOLOGY

### Methodological Framework

This investigation adopted a retrospective analytical framework, leveraging advanced ML algorithms to forecast the incidence of complicated OHSS among patients undergoing infertility treatments. The dataset for this study was meticulously composed of a wide array of patient characteristics and treatment variables as shown in Table 1.

**Table 1:**
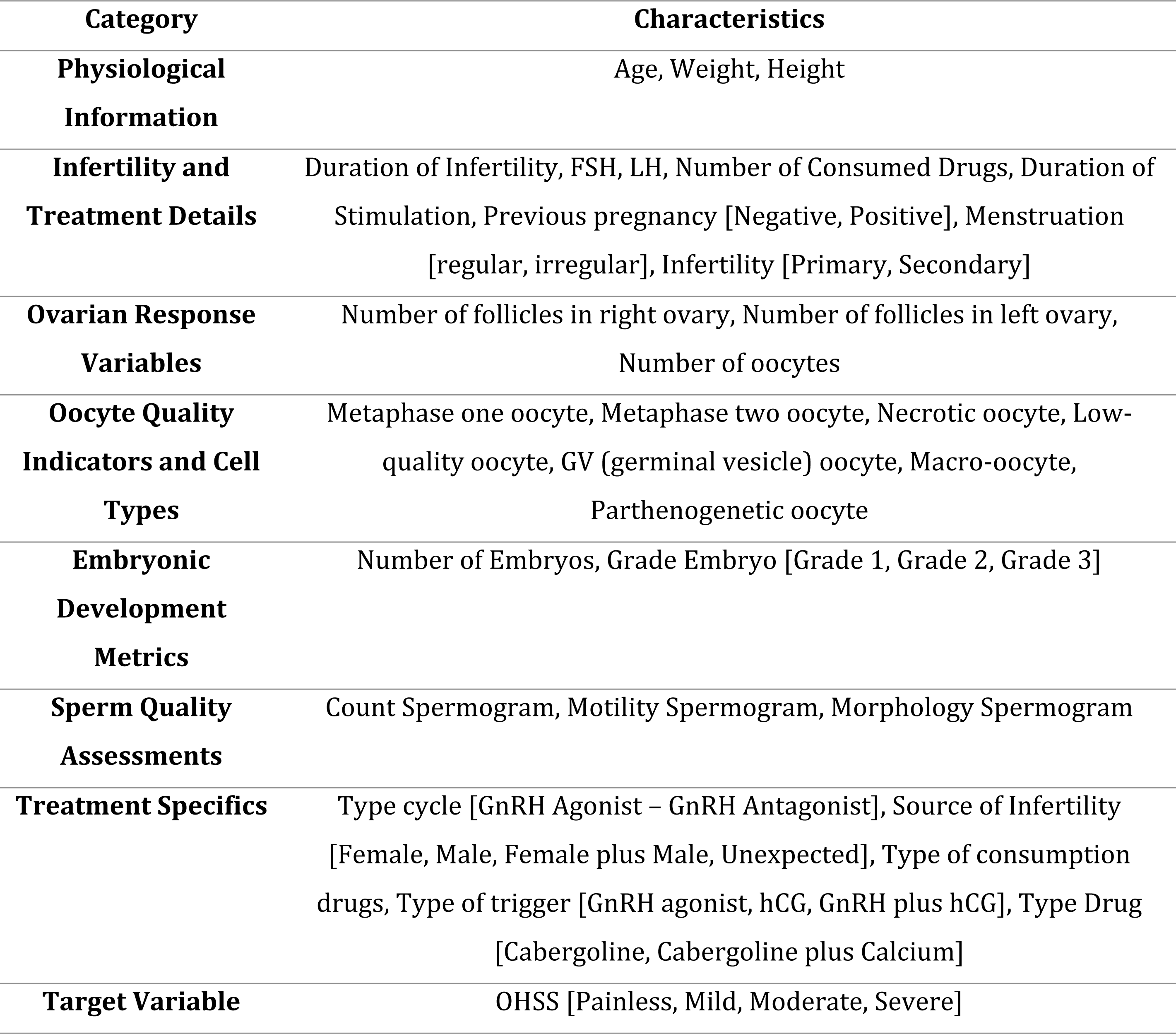
Comprehensive Overview of Parameters Used in the Study.

OHSS categories span from painless, with no significant symptoms or lab changes, to mild, moderate, severe, and critical levels, each marked by escalating severity of clinical and biochemical features. Mild OHSS manifests as abdominal discomfort and nausea without lab abnormalities, indicating its mild nature. Moderate OHSS includes these symptoms plus ascites on ultrasound, necessitating increased monitoring. Severe OHSS adds to the prior symptoms with tense ascites, severe pain, rapid weight gain, pleural effusion, and critical lab changes like elevated hematocrit and serum creatinine, indicating a significant systemic impact. Critical OHSS severely compromises vital organ functions, with potential outcomes including acute renal failure, cardiac arrhythmia, respiratory failure, thrombosis, massive hydrothorax, sepsis, and ARDS, highlighting its life-threatening nature. Our dataset did not contain critical patients.

### Development of a Machine Learning Framework

A sophisticated ML framework was engineered, incorporating Ray Tune for the nuanced tuning of hyperparameters[11]. This framework was designed to comprehensively refine the ML pipeline, covering aspects from data preprocessing and feature selection to model optimization and establishing a voting classifier system. The aim was to conduct exhaustive iterations to ascertain the most practical combination of preprocessing methodologies, algorithmic models, and their corresponding parameters.

### Ethics Statement

This retrospective study analyzed anonymized medical records with approval from the Mashhad University of Medical Sciences ethics committee (IR.MUMS.REC.1395.326). Data were accessed on July 11, 2023, and contained no identifying variables. Verbal consent was obtained from all patients for the use of their anonymized data, in accordance with ethical guidelines and data protection regulations.

### Data Preprocessing

The study aimed to forecast the occurrence of complicated OHSS. To achieve this, the target variable was transformed into a binary format: ‘painless’ and ‘mild’ cases were assigned a value of 0, while ‘moderate’ and ‘severe’ cases were labeled as 1. This binary encoding was based on the rationale that moderate and severe cases necessitate medical intervention.

To address the challenge of missing data, our preprocessing strategy employed Random Forest imputation for continuous variables and mean imputation for categorical variables, ensuring the integrity and completeness of our dataset. Additionally, we incorporated a flexible selection among various scaling options (Robust, Standard, Min-Max scalers, and a Passthrough option) and integrated the SMOTE-variants package[12, 13]. This package allowed the algorithm to choose from 60 out of 85 SMOTE variants, including oversampling and undersampling methods, to address data imbalance effectively. We ensured that extrapolated data remained within realistic boundaries by storing and applying min-max values pre-imputation. Categorical variables were processed using a 0.5 threshold for binary conversion.

### Initial Analysis

In the initial phase, multiple machine learning models were applied directly to the original dataset, which was used without any modifications for class balancing or parameter tuning. This step focused on assessing the models’ performance using all available features, considering a range of metrics. This process identified the crucial roles of data augmentation and tuning in enhancing model effectiveness.

### Dynamic Feature Selection

The feature selection process was engineered to be dynamic, allowing the search algorithm to enable or disable each variable independently. This adaptability facilitated delineating the most critical features for each model, thereby augmenting the predictive accuracy of the ML framework.

### Model Selection and Hyperparameter Optimization

The selection regime encompassed an array of models, including Decision Trees, Logistic Regression, SVM, XGBoost, LightGBM, Ridge Regression, KNN, and SGD. An extensive parameter set was provided for algorithmic determination of the most efficacious configurations for each model. The search algorithm then chose the model it saw fit and selected the required parameters. These models were subsequently integrated into a voting classifier, utilizing a soft voting mechanism to include models lacking support for predict_proba functionality.

### Objective and Evaluation Metrics

Our primary goal was to improve the recall of the positive class while ensuring that the recall for the negative class remained robust. This approach was comprehensive, considering all metrics, including recall for class 0, F1 scores, and overall accuracy. This balanced strategy was explicitly developed to navigate the challenges arising from the lack of multi-objective optimization capabilities in Ray Tune, ensuring a balanced enhancement of the model’s predictive performance.

A confusion matrix in binary classification organizes outcomes into true positives (TP), true negatives (TN), false positives (FP), and false negatives (FN), helping distinguish correct from incorrect predictions. It facilitates the calculation of key metrics: precision (accuracy of positive predictions), recall (identification accuracy within the positive class), overall accuracy, and the F1 score, harmonizing precision and recall [14, 15].

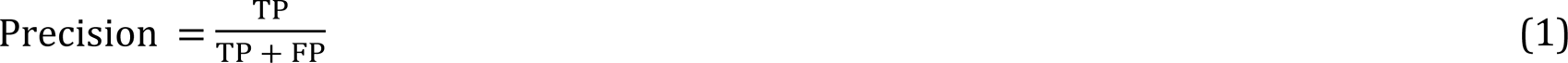

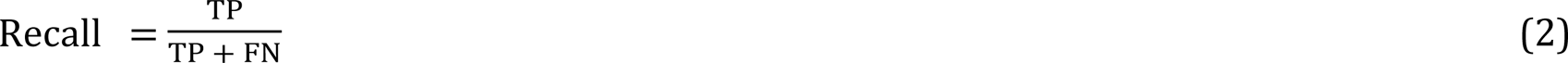

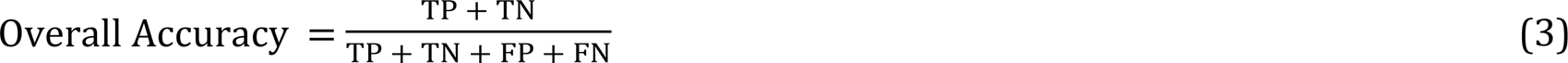

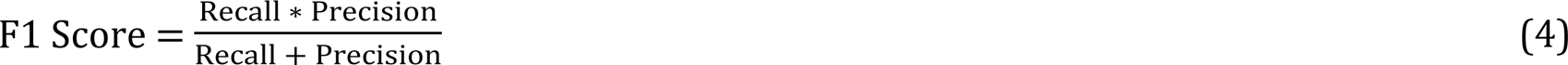

### Iterative Optimization

The optimization endeavor was structured into two distinct phases, each comprising 15,000 trials. The initial phase’s outcomes were scrutinized using the Weights and Biases package, facilitating the exclusion of underperforming models and SMOTE variants. The subsequent phase concentrated on the refined selection of models, thereby enhancing the ML framework’s efficiency and effectiveness.

### Interpretation of Models and Outcomes

The interpretative analysis of model outcomes and optimization processes was significantly bolstered by the employment of the SHAP (SHapley Additive exPlanations) package [16]. Utilizing SHAP values, which are rooted in cooperative game theory, allowed for quantifying each feature’s contribution to the predictive outcome.

## RESULT

In this section, we initiate an analysis of various developed ML models on the original data without augmentation and tuning of the models. Following this, we examine the best-performing approach, encompassing both the most effective data augmentation technique and the top-performing ML model identified in our study. Subsequently, a SHAP analysis is conducted to identify and understand the key factors contributing to the model’s predictive capabilities.

### ML models’ performance on the original data

Utilizing the methodology outlined, the ML models were initially applied to the original dataset to predict the occurrence of OHSS. Table 2 presents the results for the models, where ‘Class 1’ specifically denotes instances where the target occurs.

**Table 2:** The performance of different ML models on the real data for prediction of OHSS.

| Model/Metric | Recall<br>Class 0 | Recall<br>Class 1 | F1 Score<br>Class 0 | F1 Score<br>Class 1 | Precision<br>Class 0 | Precision<br>Class 1 | Accuracy |
| --- | --- | --- | --- | --- | --- | --- | --- |
| RandomForestClassifier | 1.00 | 0.0 | 0.95 | 0.0 | 0.90 | 0.0 | 0.90 |
| DecisionTreeClassifier | 0.96 | 0.1 | 0.93 | 0.1 | 0.91 | 0.2 | 0.87 |
| LogisticRegression | 1.00 | 0.0 | 0.95 | 0.0 | 0.90 | 0.0 | 0.90 |
| **GaussianNB** | 0.08 | 0.8 | 0.14 | 0.15 | 0.78 | 0.09 | 0.15 |
| **KNeighborsClassifier** | 1.00 | 0.0 | 0.95 | 0.0 | 0.90 | 0.0 | 0.90 |
| **SVC** | 1.00 | 0.0 | 0.95 | 0.0 | 0.90 | 0.0 | 0.90 |
| **XGBClassifier** | 0.98 | 0.1 | 0.94 | 0.15 | 0.91 | 0.33 | 0.89 |

As presented in Table 2, we explored the performance of various machine learning models in predicting complicated OHSS. These models, including RandomForestClassifier, DecisionTreeClassifier, LogisticRegression, GaussianNB, KNeighborsClassifier, SVC, and XGBClassifier, generally achieved high accuracy rates (over 85% in most cases). However, a detailed analysis uncovered a significant issue often called the ‘accuracy paradox’ in machine learning [17]. This paradox occurs when models, despite high overall accuracy, fail to effectively predict certain classes, especially the minority class, in imbalanced datasets. Specifically, while these models showed high accuracy, their performance in terms of recall and precision for class 1 (OHSS occurrence) was suboptimal. For the majority of the models, recall for class 1 was notably low (around 0 or 0.1), indicating a poor ability to identify positive OHSS cases correctly. Conversely, the GaussianNB model, although having a higher recall for class 1 (0.8), exhibited a low recall for class 0 (0.075), reflecting a skewed prediction ability. This imbalance in model performance is visually represented in Figure 1, utilizing advanced dimension reduction techniques for more precise data visualization.

**Figure 1:**
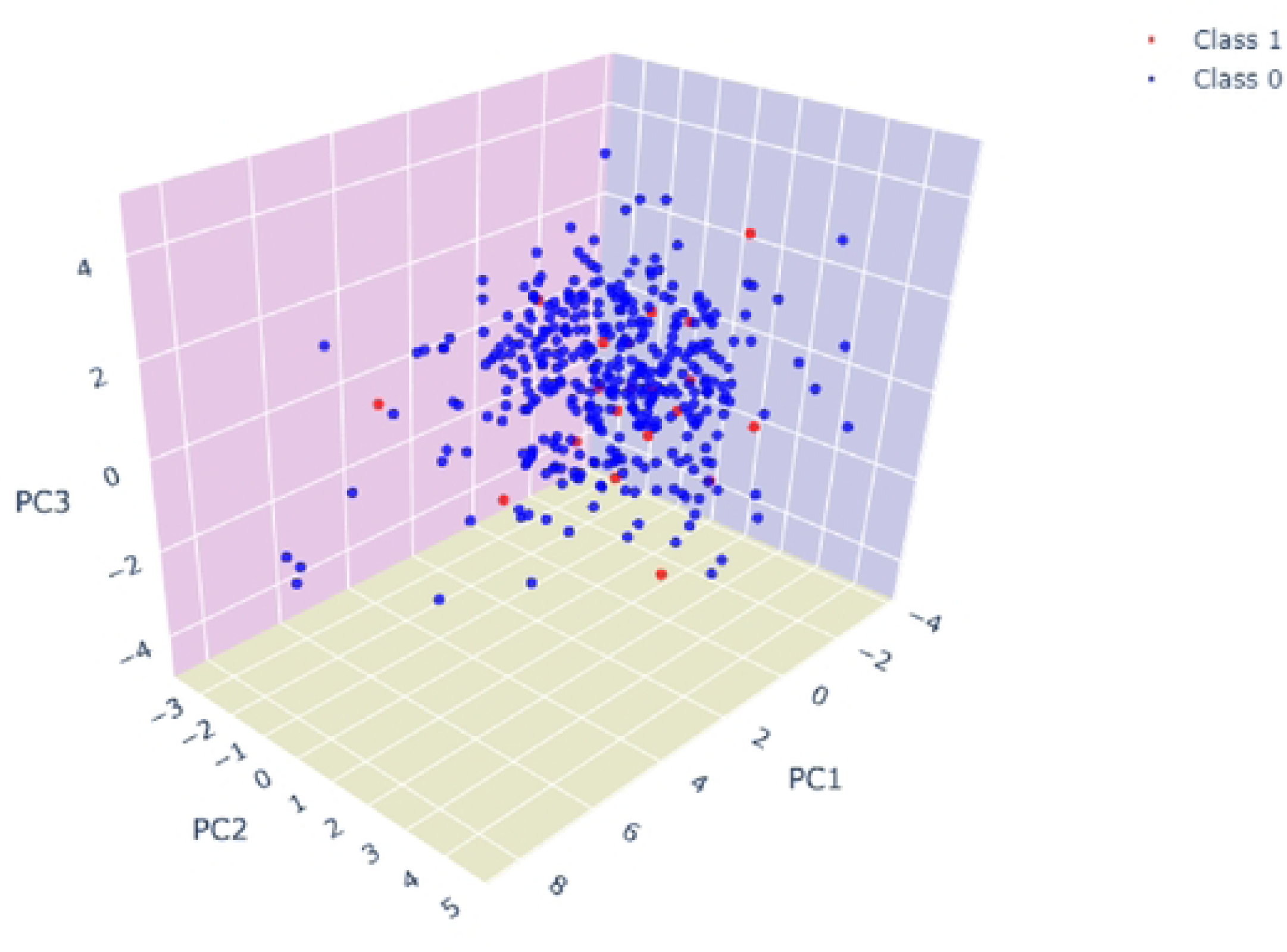
3D PCA scatter plot showcasing the distribution of two classes within the transformed feature space.

The 3D scatter plot shown in Figure 1 represents a Principal Component Analysis (PCA) output, a method commonly employed for dimensionality reduction [18]. In this plot, the data points are color-coded to represent two classes: Class 1 (red) and Class 0 (blue). The plot clearly shows an imbalance in the dataset, with Class 0 being significantly more prevalent than Class 1. Moreover, the plot reveals an overlap between Class 0 and Class 1 data points within the PCA-transformed feature space. This overlap suggests a difficulty for classifiers in distinctly separating the two classes. The classes’ proximity in this reduced dimensionality space indicates that they are not linearly separable, which could lead to decreased effectiveness in classification algorithms. This overlap and the lack of clear separation highlight the complexity of the dataset and the need for advanced or tailored ML approaches to classify such nuanced data accurately.

### Best Developed Model

As discussed in the methodology section, we analyzed different data augmentation methods coupled with different ML techniques. This approach created an ensemble learning framework, specifically using voting classifiers, which was further refined through feature tuning and hyperparameter adjustments. As a result of this extensive tuning process, a total of 15,000 distinct models were developed. Our evaluation criteria extended beyond a single metric to identify the best-performing model. We aimed for a model that demonstrated satisfactory performance across various metrics, particularly on recall for class 1. This approach ensures a more balanced and comprehensive assessment of the model’s predictive capabilities, especially in accurately identifying the minority class. Figure 2 presents the outcomes of 90 models selected from the pool of 15,000, representing the top, medium, and worst performers.

**Figure 2:**
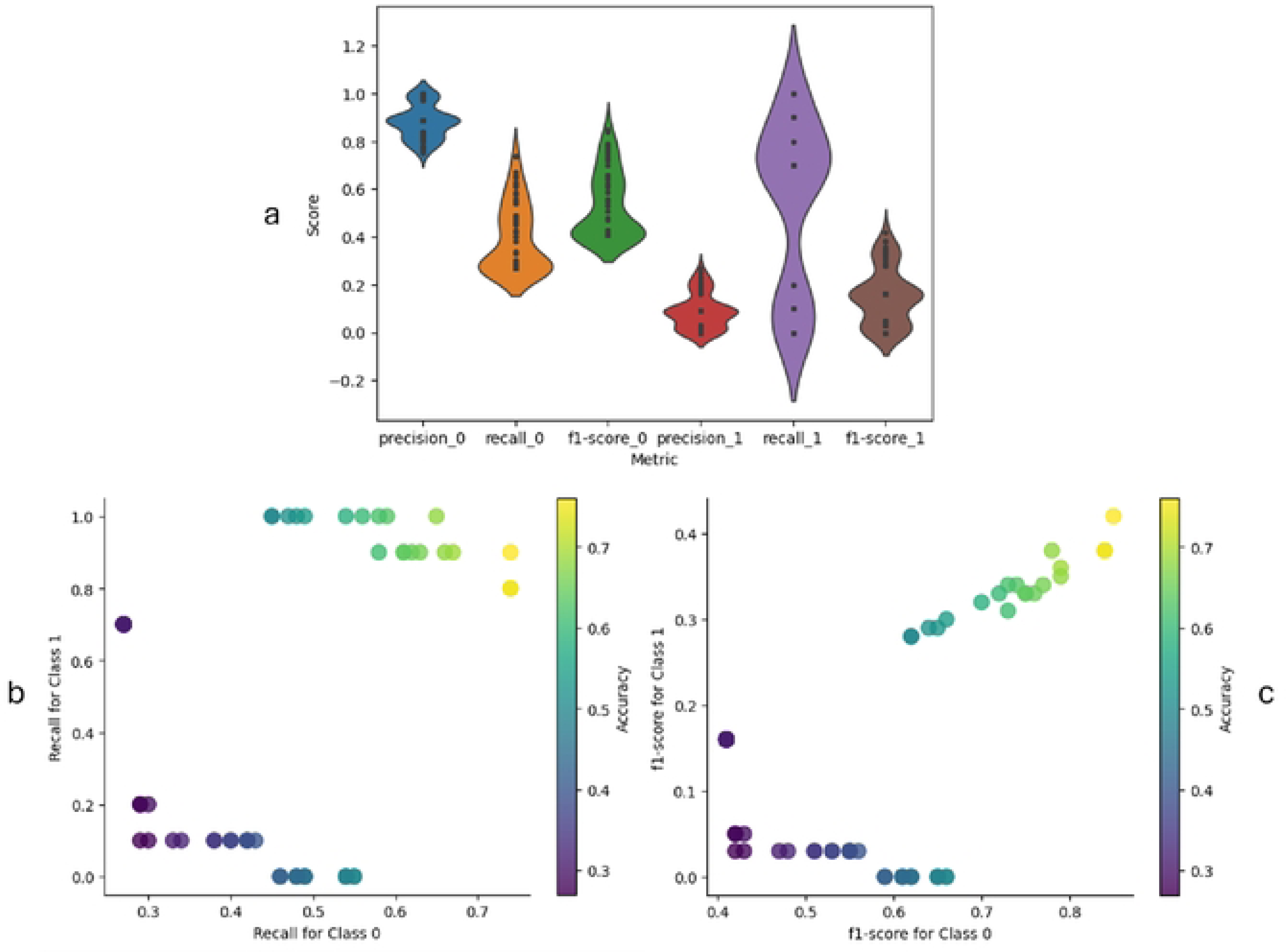
Analysis of the developed ML n1odels: a) metrics distributions, b) trade-off between recall scores, and c) trade-off between fl-scores.

Figure 2 (a) shows the distribution of key performance metrics—precision, recall, and F1-scores for both classes (0 and 1)—across the selected set of models. The variation in these metrics, especially for class 1, highlights the inherent difficulties in accurately predicting the minority class in the dataset. Figure 2 (b) demonstrates a scatter plot illustrating the trade-off between recall scores for both classes. The color gradient in this plot indicates the accuracy level of each model. Notably, the model that achieved a high recall for class 1 (0.9), which is paramount in identifying true OHSS cases, also maintained a robust recall for class 0 (0.74), thus demonstrating a balanced sensitivity across classes. In Figure 3 (c), a scatter plot features the F1-scores of both classes, with accuracy once again represented by a color gradient. The model highlighted here not only demonstrated a high recall for class 1 but also achieved the best F1 scores for both classes. This indicates an excellent balance between precision and recall, underscoring the model’s comprehensive performance capabilities.

**Figure 3:**
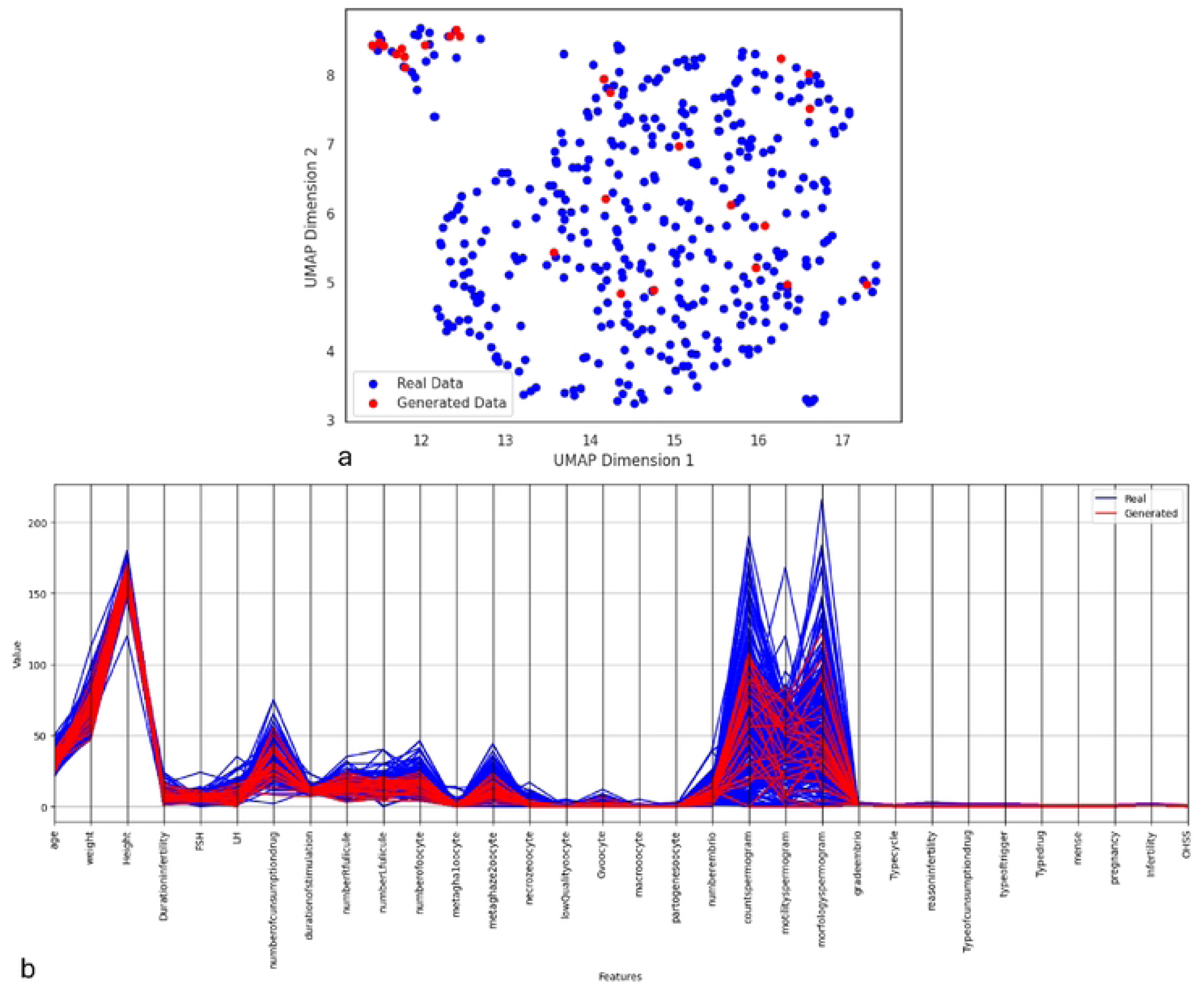
Analysis of the generated data: a) UMAP projection of the real data vs generated data, b) Parallel Coordinates plotof real vs generated Data.

In this best-developed model, the Iterative Instance Adjustment for Imbalanced Domains (IPADE-ID) algorithm [19] emerged as the most effective augmentation technique. The implementation of instance-generation techniques like IPADE-ID is increasingly acknowledged as a practical solution for addressing the challenges posed by highly imbalanced datasets. Figure 3 shows the similarity between the dataset generated using the IPADE-ID technique and the real data, highlighting the technique’s efficacy in producing representative and accurate synthetic datasets.

In Figure 3, we present a comprehensive analysis of the data generated by the IPADE-ID, aimed at mirroring the distribution of the real dataset. Figure 3 (a) features a Uniform Manifold Approximation and Projection (UMAP) visualization [20], plotting both real (blue points) and generated (red points) data in a two-dimensional space. The UMAP algorithm captures the underlying structure of high-dimensional data, revealing two distinct clusters. Notably, the synthetic data points are well-integrated with the real data points, signifying that the generative model has accurately learned the complex manifold of the real data. Figure 3 (b) offers a Parallel Coordinates plot [21], which visualizes the values of each feature across the real and generated datasets. This plot highlights the fidelity of the generated data, as shown by the overlapping lines that indicate similar feature distributions between the two datasets. The red lines, representing the generated data, closely match the blue lines of the real data across various features. This parallelism suggests that the generative model has effectively captured the real data’s central tendencies, variability, and outliers, confirming the synthetic dataset’s validity for further analysis.

Therefore, we utilized the synthetically generated dataset for the ensemble model, which was identified as the most effective. As depicted in Figure 4, the ensemble machine learning models applied to the generated datasets were SGDClassifier, SVC, and RidgeClassifier. It should be noted that when these models were not used in an ensemble configuration, they could not accurately identify any instance of OHSS occurrence, yielding a recall rate of zero for class 1. However, when applied to the data generated using the IPADE-ID technique, the ensemble model exhibited outstanding performance, particularly in predicting class 1, which indicates the occurrence of OHSS. The effectiveness of this approach is detailed in Table 3.

**Figure 4:**
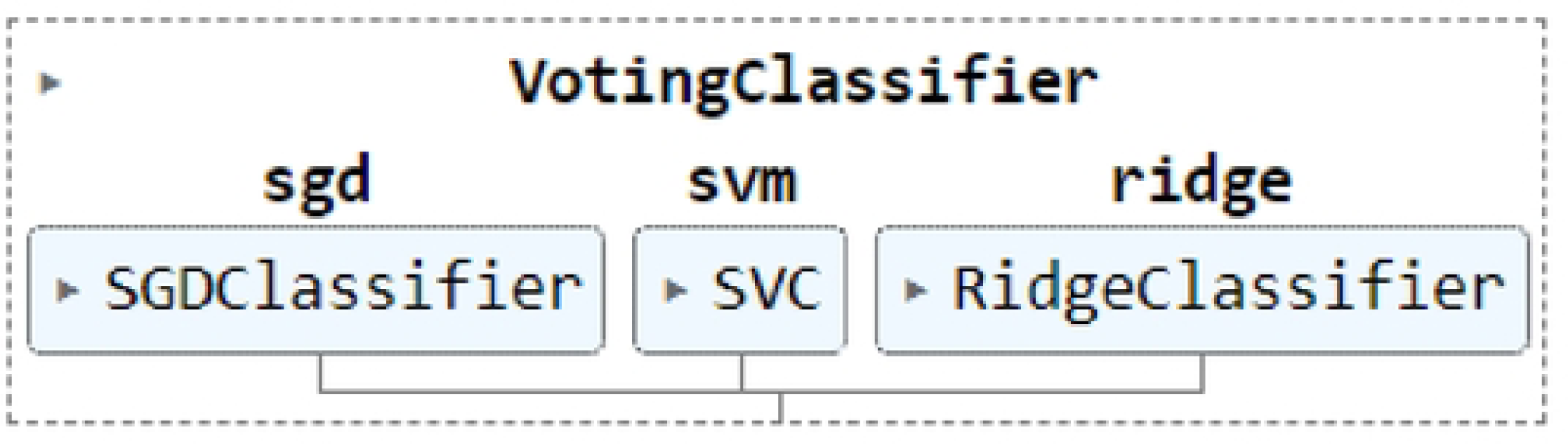
Representation of the best ensemble model.

**Table 3:**
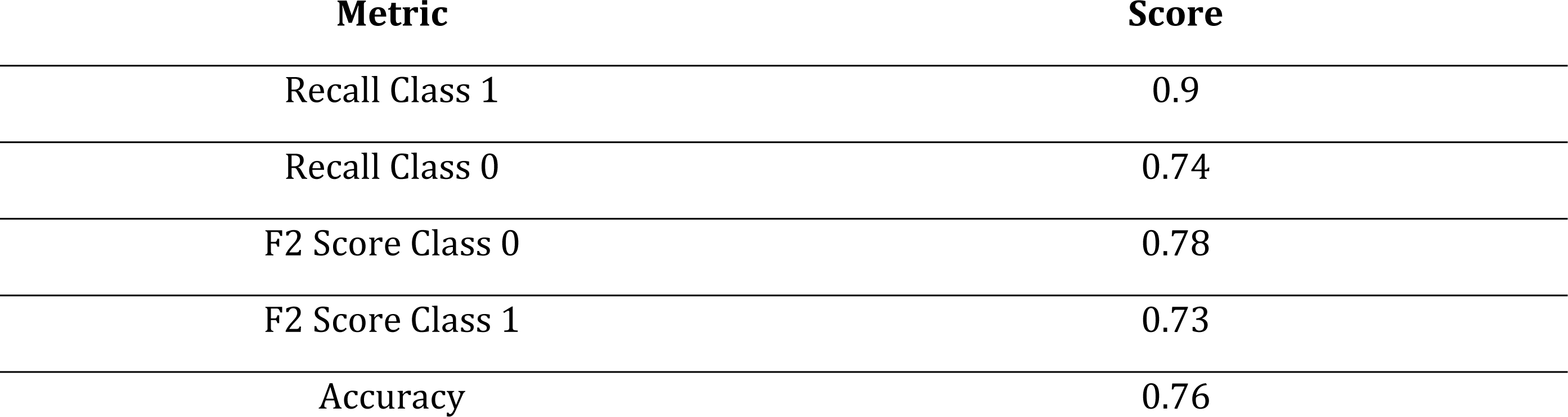
The performance of the best-developed ML model.

According to Table 2, the ensemble model accurately predicted the occurrence of OHSS 90% of the time. Additionally, the model demonstrated an overall satisfactory performance with an accuracy rate of 0.76. The model also performed well in other key metrics.

### SHAP analysis

Building on the results of the best model, the next step involves using this model to perform a SHAP analysis. This analysis helps identify and understand the most influential features in predicting OHSS occurrence, providing deeper insights into the model’s decision-making process.

Figure 5 presents a SHAP summary plot, providing a visual representation of how the selected features from the tuning phase influence the model’s output. On the X-axis, the SHAP values quantify the impact of each feature, with zero indicating no impact and higher absolute values (positive or negative) indicating a more significant influence on the model’s predictions. The Y-axis lists the features used in the model, and the color represents the value of the features, where one side of the color spectrum (blue) shows a low feature value, and the other side (red) shows a high feature value. Key features such as “Typecycle” and “reasoninfertility” stand out in the plot, signifying their crucial effect on determining the model’s output. A closer examination reveals those higher values of “reasoninfertility” correlate with a decreased likelihood of predicting OHSS, representing the risk of complicated OHSS decreases in cases of solely male infertility or combined male and female infertility since this feature mostly aligns with the negative side of the SHAP value spectrum. Conversely, elevated values in “Typecycle” and “mense” [irregular or irregular menstruation cycles], which denote the use of GnRH agonist and the presence of irregular menstruation, respectively, positively impact the model’s output. These factors are associated with an increased probability of accurately predicting OHSS. These findings are extensively analyzed and discussed in the discussion section of our paper, where we delve deeper into how these features collectively influence the model’s predictions and the implications of their respective impacts on the risk of OHSS.

**Figure 5:**
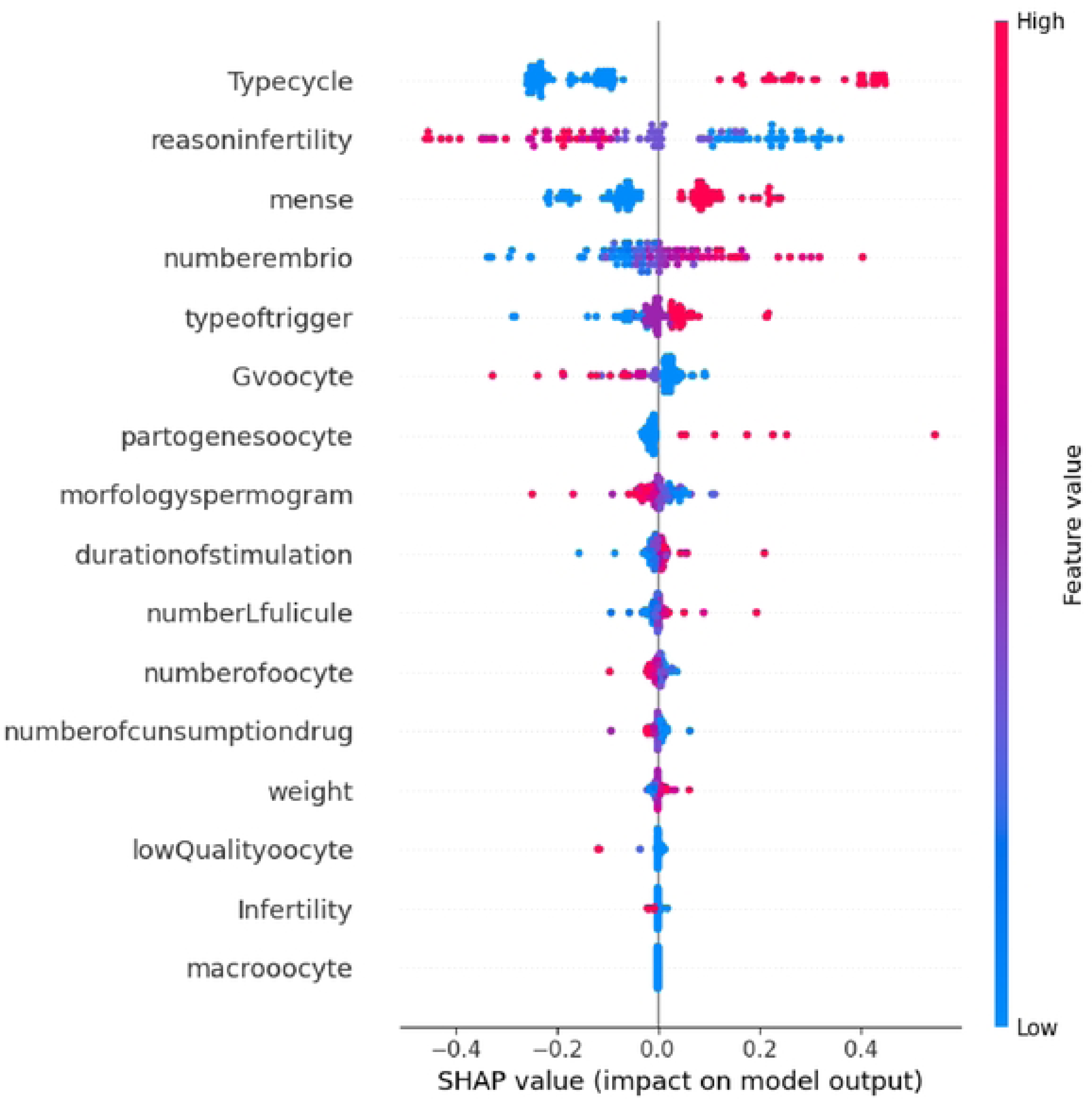
SHAP analysis of the ensembled model for prediction of OHSS.

## DISCUSSION

This study represents a pioneering effort in using ML to predict complicated OHSS, addressing a gap in the existing literature. Confronting the challenge of a highly imbalanced dataset, we employed data augmentation techniques and optimized a voting classifier’s hyperparameters, achieving notable results. Our work diverges from traditional infertility research, which often focuses on predicting successful pregnancy or oocyte retrieval outcomes, by specifically addressing the less-explored complications of infertility treatments like OHSS. This approach not only fills a significant gap in the literature but also provides new perspectives on the prevention and management of such infertility treatment complications. Table 4 displays a comprehensive comparison of our study’s contributions with similar research in the literature, highlighting how this research advances and enhances

**Table 4.**
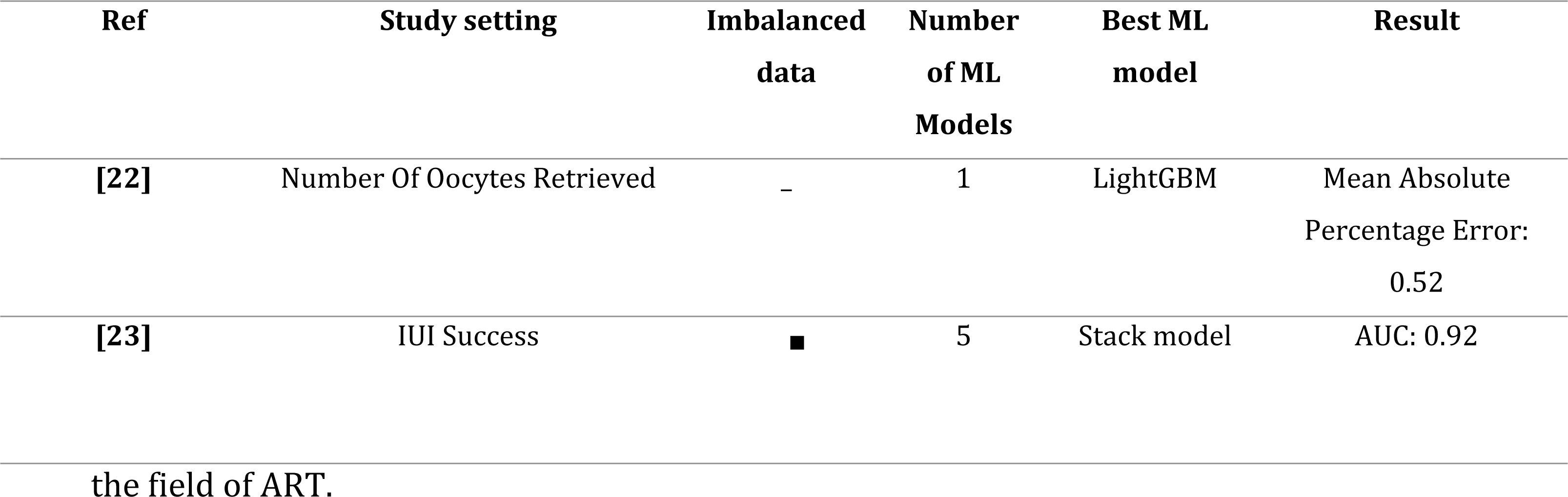
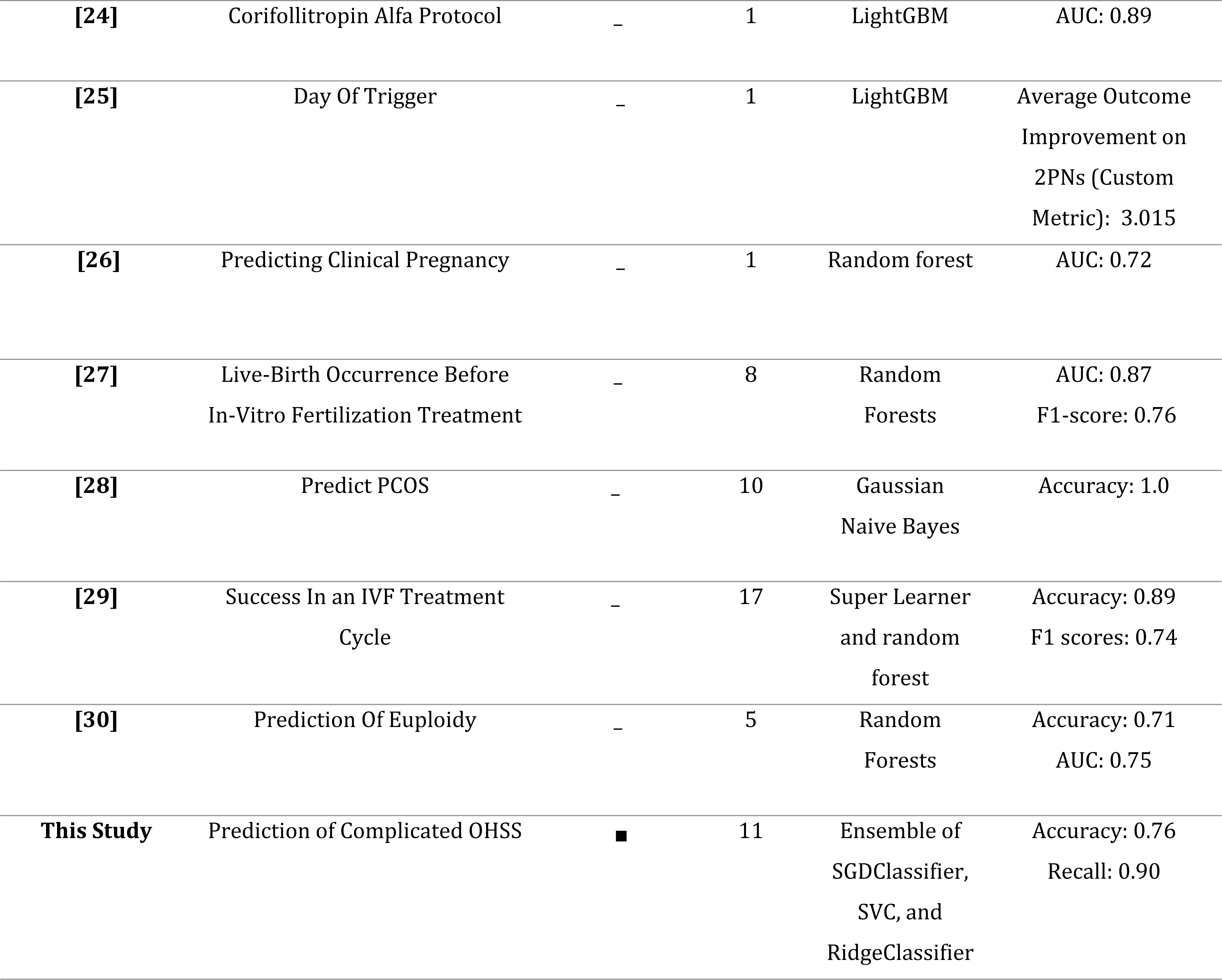
Similar Studies That Have Utilized ML in the Field of ART.

| Ref | Study setting | Imbalanced data | Number of ML Models | Best ML model | Result |
| --- | --- | --- | --- | --- | --- |
| [22] | Number Of Oocytes Retrieved | – | 1 | LightGBM | Mean Absolute Percentage Error: 0.52 |
| [23] | IUI Success | ■ | 5 | Stack model | AUC: 0.92 |

Table 4. Similar Studies That Have Utilized ML in the Field of ART
|  |  |  |  |  |  |
| --- | --- | --- | --- | --- | --- |
| [24] | Corifollitropin Alfa Protocol | – | 1 | LightGBM | AUC: 0.89 |
| [25] | Day Of Trigger | – | 1 | LightGBM | Average Outcome Improvement on 2PNs (Custom Metric): 3.015 |
| [26] | Predicting Clinical Pregnancy | – | 1 | Random forest | AUC: 0.72 |
| [27] | Live-Birth Occurrence Before In-Vitro Fertilization Treatment | – | 8 | Random Forests | AUC: 0.87<br>F1-score: 0.76 |
| [28] | Predict PCOS | – | 10 | Gaussian Naive Bayes | Accuracy: 1.0 |
| [29] | Success In an IVF Treatment Cycle | – | 17 | Super Learner and random forest | Accuracy: 0.89<br>F1 scores: 0.74 |
| [30] | Prediction Of Euploidy | – | 5 | Random Forests | Accuracy: 0.71<br>AUC: 0.75 |
| **This Study** | Prediction of Complicated OHSS | ■ | 11 | Ensemble of SGDClassifier, SVC, and RidgeClassifier | Accuracy: 0.76<br>Recall: 0.90 |

### GnRH Antagonists vs. GnRH Agonists and Stimulation Duration

The results of our machine learning study using SHAP revealed that the use of GnRH antagonists, compared to GnRH agonists, and longer stimulation duration were identified as important factors in predicting the occurrence of complicated OHSS. The existing literature largely supports these findings. A comprehensive Cochrane review provided moderate quality evidence that GnRH antagonists significantly reduce the incidence of OHSS without compromising live birth rates compared to long-course GnRH agonist protocols [30]. Similarly, a clinical trial focusing on PCOS patients found that the GnRH antagonist group had a significantly lower incidence of OHSS while maintaining similar pregnancy rates compared to the GnRH agonist group [31]. A large RCT and a prospective, randomized study also reported a significantly lower incidence of OHSS in the GnRH antagonist group compared to the GnRH agonist group while maintaining similar ongoing pregnancy rates (OPR) and live birth rates in both protocols [32–34]. These findings, taken together with the results of our machine learning study, suggest that the use of GnRH antagonists is an important factor in predicting and potentially reducing the occurrence of complicated OHSS. The identification of longer stimulation duration as a predictive factor for OHSS also highlights the need for further research to optimize stimulation protocols and minimize the risk of this potentially life-threatening complication.

### Reason [Source] of Infertility

Our ML model, using SHAP, found that the source of infertility is a significant predictor for complicated OHSS. The risk of complicated OHSS increases when the female factor is more involved, while the risk decreases in cases of solely male infertility or combined male and female infertility. This finding can be attributed to the fact that female-factor infertility often requires more aggressive ovarian stimulation protocols to overcome underlying issues such as ovulatory disorders or poor ovarian reserve [35]. These aggressive stimulation protocols, involving high doses of gonadotropins, are a crucial risk factor for developing OHSS. Additionally, certain genetic predispositions and biochemical factors, such as young age, lower time of infertility, lower baseline FSH, higher female factor and PCOS phenotype, body mass index, and age, and a high number of follicles and elevated estradiol levels, are associated with a higher risk of developing OHSS [35]. In contrast, when infertility is primarily due to male factors or unexpected or both female and male, the female partner may undergo less aggressive stimulation protocols, especially when using techniques like ICSI, where only a few eggs are needed, thus reducing the risk of OHSS.

### Types Of Women Infertility, Irregular vs. Regular Menstrual Cycles, Weight And PCOS

Our model identified irregular menstruation cycles, higher weight, and primary infertility as important predictors of complicated OHSS occurrence, which broadly aligns with the literature. Irregular menstruation can be associated with conditions that cause hormonal imbalances, such as PCOS, a condition known for its increased risk of OHSS[36–38]. However, the relationship between weight and OHSS remains inconclusive, with some studies finding no significant correlation [39] and others suggesting that obesity may decrease the risk of OHSS-related complications[40]. Interestingly, the model assigned lower importance to secondary infertility as a predictor of OHSS, which contradicts previous findings linking conditions that cause secondary female infertility, such as pituitary adenoma, to an increased incidence of OHSS[41]. These discrepancies highlight the need for further research to clarify the role of weight and secondary infertility in complicated OHSS prediction.

### Number of Oocytes, Embryos, Other Cell Types, and Rule of Sperm Morphology

Our machine learning model for predicting complicated OHSS presents novel insights that diverge from conventional wisdom and prior research findings. Contrary to established risk factors such as the number of retrieved oocytes, which studies like those by Verwoerd et al. suggest increase OHSS risk significantly beyond a 15-oocyte threshold[42, 43], our model de-emphasizes the importance of oocyte count. Instead, it identifies a surprising correlation between the number of embryos and higher complicated OHSS risk, assigning significant weight to the presence of 11 or more embryos as a critical predictor. This finding challenges the traditional understanding, as higher oocyte retrieval numbers have been linked to increased OHSS risk without corresponding benefits in live birth rates in fresh autologous IVF cycles. Our analysis also highlights the potential predictive value of other cell types, including pathogenetic oocytes, GV oocytes, and macroocytes for complicated OHSS cases. Furthermore, while no direct studies have linked sperm morphology with OHSS incidence, our observations suggest that improved sperm quality may contribute to better-fertilized oocyte outcomes, indirectly influencing complicated OHSS risk. This aligns with the literature on the predictive value of normal sperm morphology for pregnancy success in IUI and IVF, where a significant improvement in pregnancy rates was noted for strict sperm morphology criteria[44].

### Types of Triggers

The utilization of different trigger drugs in ovarian stimulation protocols has been shown to influence the risk of complicated OHSS. Our model, using SHAP analysis, found that the use of hCG or hCG combined with GnRH agonists as trigger drugs significantly increases the risk of complicated OHSS. In contrast, the use of GnRH agonists alone was a predictor of uncomplicated OHSS. These findings align with previous literature, which has shown that GnRH agonists result in a significantly lower incidence of moderate to severe OHSS compared to hCG, albeit with lower live birth and ongoing pregnancy rates[45, 46]. While dual trigger with GnRH agonists and low-dose hCG may be associated with a modest increase in oocyte yield[47], the risk of OHSS following dual triggering remains unclear[48]. A recent study recommends the use of a single GnRH agonist trigger for high responders treated with the freeze-all strategy to prevent moderate to severe OHSS and obtain satisfactory pregnancy and neonatal outcomes in subsequent frozen embryo transfer cycles[49].

Several key limitations, including the reliance on a small dataset from a single center, can affect the generalizability of our findings. Additionally, the absence of an early prediction mechanism and the study’s non-utilization of time series analysis limit our model’s ability to preemptively identify OHSS risk and dynamically track its progression over time. These constraints highlight areas for future research to enhance predictive accuracy and clinical utility.

## CONCLUSION

In conclusion, our pioneering study demonstrates the potential of machine learning models in predicting complicated OHSS, a previously unexplored area in infertility research. Despite the challenges posed by the highly imbalanced dataset, we successfully implemented various data augmentation techniques and optimized the hyperparameters of a voting classifier to achieve satisfactory results. Our findings not only contribute to the growing body of research on machine learning applications in assisted reproductive technology but also provide valuable insights into the factors influencing the occurrence of complicated OHSS.

## AUTHOR CONTRIBUTIONS

A.Z. and H.K. developed the machine learning framework, performed data analysis, and revised the drafted manuscript [Software], [Formal Analysis], [Writing – review & editing]. M.M. was responsible for data acquisition, supervision, and preparation of the final draft [Data curation], [Supervision], [Writing – original draft]. I.A. contributed to the conceptualization, supervision of the study and final draft preparation [Conceptualization], [Supervision], [Writing – original draft]. T.S. prepared the original draft and assisted in data acquisition [Writing – original draft], [Data curation]. All authors contributed to the evaluation and interpretation of the results [Validation]. All authors approve of the submitted manuscript.

## DATA ACQUISITION AND ETHICAL CONSIDERATIONS

Data collection adhered strictly to ethical standards, with approval from the relevant university ethics committee with approval code of IR.MUMS.REC.1395.326. Before inclusion in the study, consent was obtained from all patients to use their anonymized data for research and analysis purposes.

## ACKNOWLEDGMENTS

We want to extend our gratitude to Prof. Mehdi Jabbari Nooghabi, whose guidance and insights have been instrumental in the development of this machine learning framework.

## CONFLICT OF INTEREST

The authors declare no potential conflicts of interest with respect to the research, authorship, and/or publication of this article.

## FUNDING

This research received no specific grant from any funding agency in the public, commercial, or not-for-profit sectors.

## DATA AVAILABILITY STATEMENT

The dataset supporting the conclusions of this article is confidential and cannot be made publicly available. However, data may be available from the corresponding author upon reasonable request and with permission of the original data providers, subject to compliance with applicable confidentiality agreements and data protection laws.

### Declaration of Generative AI and AI-assisted Technologies in the Writing Process

In the development and composition of this document, generative AI was not utilized beyond the scope of fundamental tools for grammar, spelling, and reference verification.

### Summary Table

What was already known on the topic:

- Machine learning models have been applied to predict outcomes like oocyte retrieval numbers, IUI success, pregnancy rates, and live birth rates in assisted reproductive technology (ART)
- Ovarian hyperstimulation syndrome (OHSS) is a potentially life-threatening complication of ART, but prediction of complicated OHSS using machine learning has not been explored previously
- Imbalanced datasets are a major challenge for developing effective machine learning models in medical domains

What this study added to our knowledge:

- This is the first study to develop and optimize machine learning models specifically for predicting complicated cases of OHSS, addressing the obstacle of imbalanced data
- The optimized ensemble model provides insights that challenge certain conventional assumptions about risk factors for OHSS, such as de-emphasizing oocyte numbers while highlighting the number of embryos as a predictor
- Novel data augmentation techniques like IPADE-ID were effectively applied to tackle the highly imbalanced nature of the OHSS dataset
- The study identified key influential factors like GnRH antagonist use, stimulation duration, female infertility factors, irregular menses, higher weight, hCG trigger usage, and number of embryos through interpretation of the optimized model

## Notes

### Competing Interest Statement

The authors have declared no competing interest.

### Funding Statement

The author(s) received no specific funding for this work.

